# Driver gene detection via causal inference on single cell embeddings

**DOI:** 10.1101/2024.05.16.24307521

**Authors:** Chengbo Fu, Lu Cheng

## Abstract

Driver genes are pivotal in different biological processes. Current methods generally identify driver genes by associative analysis. Leveraging on the development of current large language models (LLM) in single cell genomics, we propose a causal inference based approach called CID to identify driver genes from scRNA-seq data. Through experiments on three different datasets, we show that CID can (1) identify biologically meaningful driver genes that have not been captured by current associative-analysis based methods, and (2) accurately predict the change directions of target genes if a driver gene is knocked out.

## 1 Introduction

In cancer biology, a driver gene is defined as a gene that significantly affects the expression of downstream genes, altering cellular behavior and contributing to the malignant phenotype. Therefore, driver genes play critical roles in cancer development. However, identifying driver genes is not an easy task.

Traditional methods identify potential driver genes in cancer mainly based on DNA-seq or RNA-seq data. The DNA-seq based approaches, such as MutSigCV [1], OncodriveFM [2], and OncodriveCLUST [3], are based on the idea that cancer cells try to gain survival advantage over normal cells by mutating the driver genes, so the mutation frequencies of driver genes in the cancer tissue are higher than that in the normal tissue. The RNA-seq based approaches, such as CONEXIC [4] and TieDIE [5], are based on the idea that potential driver genes are differentially expressed in the cancer tissue, so differential gene expression (DE) analysis could be used for potential driver gene detection.

Differentially expressed genes may not be real driver genes because (1) they may be downstream genes of the driver gene and (2) the driver gene may not necessarily be highly differentially expressed as a small change of driver gene is enough to cause the malignant phenotype, e.g. transcription factors. Therefore, real driver gene identification relies on further wet lab experiments, which are laborious and expensive to perform. To validate a driver gene, researchers usually knock out (KO) or overexpress a potential driver gene in cancer cell lines to see if the malignant phenotype disappears or is enhanced. Over the years, a manually curated database NCG (Network of Cancer Genes) [6] has collected cancer driver genes reported in the literature, which could be used as a reference for potential driver genes.

As we are entering the era of single cell genomics, it is natural to identify potential driver genes using DE analysis on single cell RNA-seq (scRNA-seq) data. The advantage of scRNA-seq is that we could classify cells into different cell types to decipher the tissue heterogeneity and perform differential analysis between cell types. The differentially expressed driver genes are called marker genes. Since we are now comparing the cell types and scRNA-seq data may not be generated from cancer tissues, the concept of cancer driver gene is not defined. However, we want to borrow the term “**driver gene**” and use it to refer to genes that affect the expression of lots of downstream genes and determine cell type differentiation. Note that the driver genes are not equivalent to marker genes that are generated by DE analysis.

There exists several limitations if we directly use marker genes derived from scRNA-seq data as driver genes. First, marker genes are derived by DE analysis, which is associative rather than causal. As a result, the marker genes may be the downstream genes of driver genes, while driver genes might not appear on the top of the DE gene list. Another limitation is that validation experiments of the potential driver genes are laborious and expensive. It will significantly reduce the costs if we could perform in silico validation of the potential driver genes.

Current developments of large language models (LLM) on scRNA-seq data such as scBERT [7], scGPT [8] have provided a potential solution to address the aforementioned limitations. LLMs are trained on a diverse range of scRNA-seq datasets to provide cell and gene embeddings in a common embedding space. We could mask a set of genes from the input and ask LLMs to predict their embeddings, which could be utilized to study the relationships between different genes, e.g. identifying driver genes.

Current LLMs serve as the foundation models rather than directly solve the driver gene identification problem. scBERT uses attention scores to find marker genes, which is an associative analysis. The attention score depicts the correlation between two genes. Causal analysis, on the contrary, considers the direction between two genes. The impact of gene A to gene B is not the same as vice versa. However, we have not been able to find such a causal analysis based approach from the literature that addresses the driver gene identification problem.

To fill the gap, we propose a Causal inference approach to Identify Driver genes (CID) by leveraging the power of LLM scBERT. Let us illustrate the idea using an example where we want to study the impact of gene A to gene B. First, we will mask gene B from an input cell and predict its embedding *e*_1_. Next we mask both gene A and B from the input cell and again predict gene B’s embedding *e*_2_. If gene A is an upstream gene, i.e. driver gene, then the second embedding *e*_2_ will be far away from the first embedding *e*_1_ as gene A has a huge impact on gene B; otherwise the two embeddings will be very close as gene A has little impact to gene B. The operation of masking gene A and B together mimics the *do*(*X*) operation in causal inference, i.e. the intervention.

We demonstrate CID’s new insights on three datasets. We first illustrate that CID can identify new driver genes with biological interpretations in delta pancreas islet cells, which are not picked by scBERT. Then, we show CID accurately predicts the impact of driver genes towards their target genes in a perturb-seq data consisting of 19 knockout experiments. Finally, we compare CID with a recent driver gene identification method CSDGI on a scRNA-seq data of breast cancer, where CID has identified more driver genes (*n* = 12) that have annotations in the driver gene database NCG 7.1 [6].

## 2 Data and Methods

Pancreas islet cells scRNA-seq data (Muraro dataset) was downloaded from Gene Expression Omnibus (GEO) GSE85241. Perturb-seq datasets (Dixit dataset) was downloadded from GEO GSE90063. Breast cancer data was downloaded from GEO GSE75688.

All downloaded data were provided in the form of a gene expression matrix. We preprocessed the gene expression matrices following the same steps as scBERT. The expression of each cell is mapped to a 16,906-dimensional vector, with each dimension representing a unique human gene. Log-normalization is performed on the data using a size factor of 10,000. Cells with fewer than 200 expressed genes are filtered out. By default, the pre-trained weights of scBERT are used in CID analysis.

All DE analysis were performed using Scanpy (v1.9.8) [9] with the `sc.tl.rank_genes_groups` function (`wilcoxon` as the test method).

In pancreas islet cells analysis, we pooled all the marker genes identified by scBERT both as the potential driver genes and target genes in CID analysis. In case of calculating the impact of a driver gene to itself, we directly set it to 0. For each cell type, we take the top 10 driver genes and compare it with scBERT.

The perturb-seq data (Dixit dataset) contains 19 driver gene knock-out (KO) and wild-type (WT) pairs. After DE analysis, we filter genes that have at least 20 reads in both KO and WT samples for downstream analysis. Up-regulated genes are selected by *pvalue* < 0.05 and *log*2*fc* > 1. Down-regulated genes are *pvalue* < 0.05 and *log*2*fc* < − 1. No-change genes are |*log*2*fc*| < 0.5. From each category, 10 target genes are uniformly sampled for CID prediction. The `nn.softmax` function in `predict.py` of scBERT was used to convert embeddings into discrete gene expression levels.

In breast cancer data analysis, we feed the gene expresion matrix to the scBERT script `pretrain.py` with default parameters to re-train scBERT. Cells are divided into cancer and normal cells based on the `information.csv` file.

## 3 Results

CID performs causal inference based on the framework of scBERT. We denote the input scRNA-seq gene expression matrix by *X* = (***x***_1_, ***x***_2_, …, ***x***_*N*_), where ***x***_*i*_ = (*x*_*i*1_, *x*_*i*2_, …, *x*_*iG*_) denotes the expression profile of the *i*-th cell and *x*_*ij*_ ∈ {0, 1, 2, 3, 4, #} denotes the expression level of *j*-th gene in the *i*-th cell. Note that scBERT categorizes the continuous gene expression levels into 5 discrete levels and # is reserved for the masking operation.

The scBERT network, denoted by *f*_scBERT_, transforms the gene expression matrix *X* into a latent space ***Z*** = (***z***_1_, ***z***_2_, …, ***z***_*N*_):

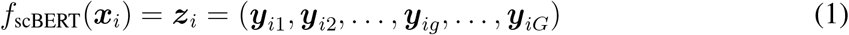

where ***y***_*ig*_ = (*y*_*ig*1_, *y*_*ig*2_, …, *y*_*igD*_) is a D-dimensional embedding for the *g*-th gene in the *i*-th cell and ***z***_*i*_ is a concatenated vector of ***y***_*ig*_ for *g* = 1, 2, …, *G*.

To quantify the impact of gene *j* to gene *k*, we introduce the mask notations 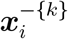 and 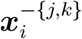, which refers to masking the gene expression level of gene sets {*k*} and {*j, k*}, respectively. 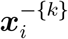 and 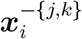 are defined as

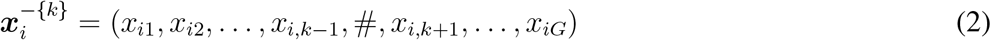

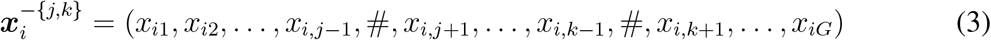

The predicted embeddings of scBERT based on 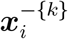 and 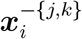 are given by

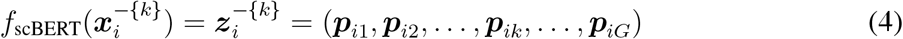

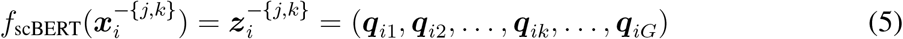

Therefore, the impact score of gene *j* to gene *k* in cell *i* is given by the Euclidean distance ∥***p***_*ik*_ − ***q***_*ik*_ ∥ between the two predicted embeddings of gene *k*. The average impact of gene *j* to gene *k* over all *N* cells is given by

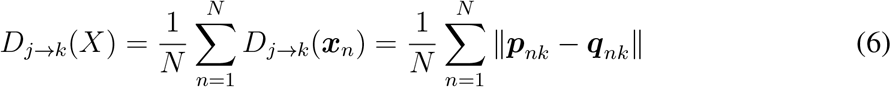

In real data analysis, due to the high computational load, we have to select a set of potential driver gene ***s*** = (*s*_1_, *s*_2_, …, *s*_*r*_, …, *s*_*R*_) and a set of target genes ***t*** = (*t*_1_, *t*_2_, …, *t*_*m*_, …, *t*_*M*_), where *s*_*r*_, *t*_*m*_ ∈ {1, 2, …, *G*}. Usually the potential driver genes and target genes are chosen as DE genes with a relaxed threshold. We will compute the following marginal impact for each driver gene *s*_*r*_ over all target genes ***t*** given by

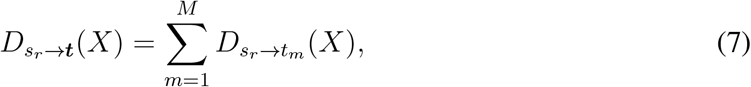

where 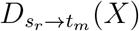 is calculated by Eq. 6. The potential driver genes are then ranked by their marginal impact (Eq. 7) and used as the final result of CID. The higher the impact, the more likely the gene is a driver gene.

Fig. 1A shows the general workflow of CID. For each target gene, we derive its impact to all target genes, from which we calculate its marginal impact. We rank the potential driver genes by their marginal impacts and select the top ones as the final candidate for wet lab validation. As illustrated in Fig. 1B, the driver gene A have large impact scores to downstream genes B and C, while the impact score of B to C is relatively small as B is not a parent of C.

**Figure 1:**
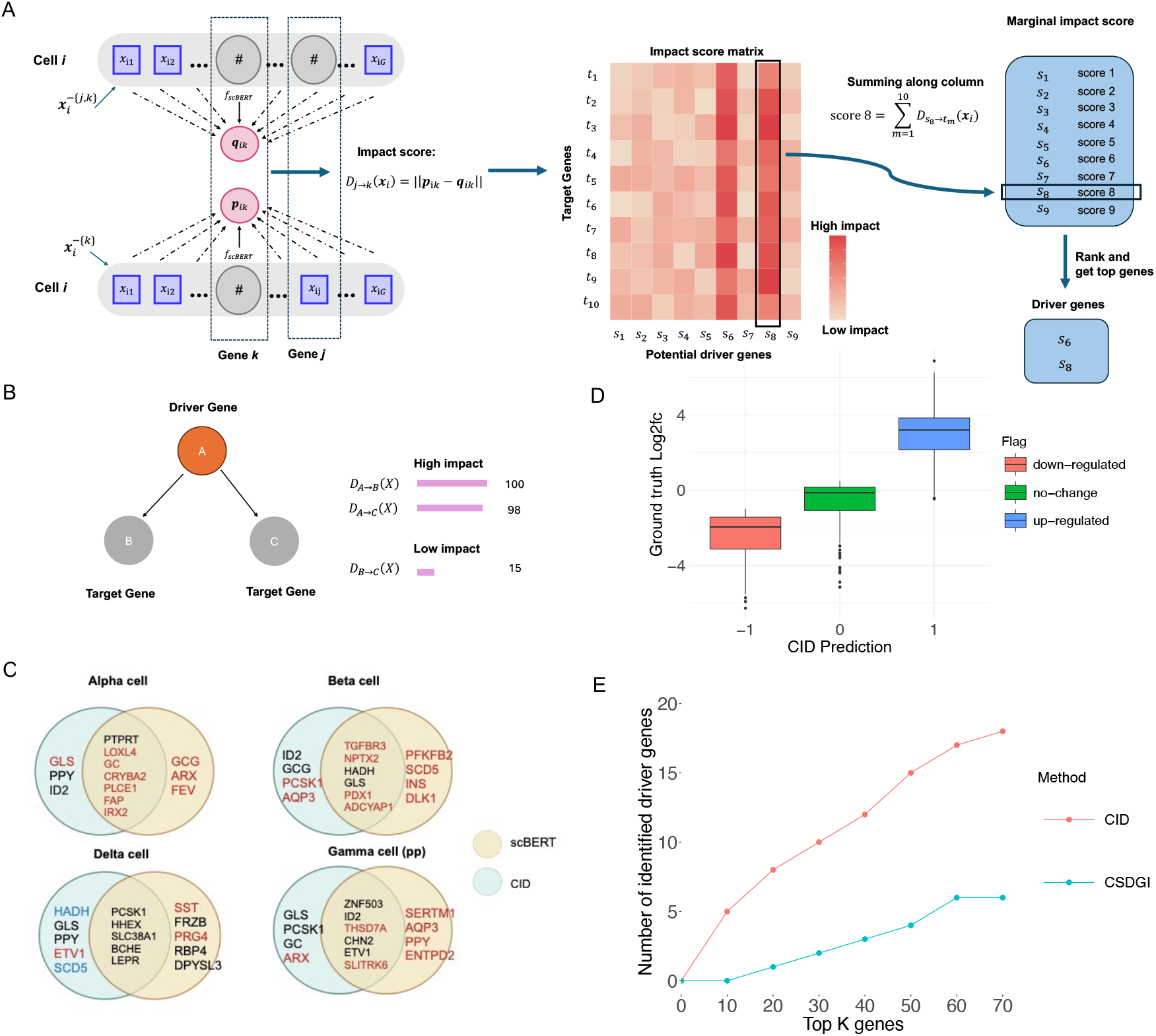
(A) CID workflow. (B) Toy example of driver gene. (C) CID vs scBERT predicted driver genes. Red and blue fonts indicate database (CellMarker 2.0) annotated marker genes and literature-supported driver genes. (D) CID predicted gene expression change vs ground truth log2fc. (E) CID vs CSDGI in driver gene identification.

### 3.1 CID vs scBERT on Pancreas islet cells data

We applied CID to a scRNA-seq dataset of pancreas islet cells that had been analysed by scBERT, where gene expression matrix and cell type annotation were provided. Following the analysis of scBERT, we focused on the driver gene detection on 4 cell types: alpha cells, beta cells, delta cells and gamma cells. The gene expression matrices of the four cell types were fed into CID independently to identify cell-type specific driver genes. Fig. 1C shows the Venn diagram between the driver genes of CID and the marker genes given by scBERT based on attention scores, where genes in red color are marker genes given by CellMarker 2.0 database [10]. It is obvious that scBERT finds more marker genes than CID, which suggests the attention score approach in scBERT is an associative analysis.

CID identified multiple potential driver genes that were not marker genes and not found by scBERT in different cell types. We performed a survey of the literature of these genes and found that HADH and SCD5 in delta cells (blue fonts in Fig. 1C) were reported as driver genes in previous publications [11, 12]. HADH is reported to cause monogenic diabetic cell disorders via delta cells [11]. SCD5 is reported [12] to be highly expressed in delta cells and regulate downstream transcription factors such as SOX9, MYC and HES1, which are important for cell differentiation. These reports suggest that CID identifies biologically meaningful driver genes, which are missed by scBERT.

### 3.2 Validation of CID prediction on perturb-seq data

We validate the up-regulation and down-regulation of target genes based on CID using perturb-seq data. The perturb-seq data is generated from 19 experiments, in each of which a driver gene is knocked out using CRISPR-Cas9 in K562 cells. scRNA-seq data is generated for the cells before (WT) and after (KO) the driver gene knock-out.

Based on the gene expression matrix of WT K562 cells, i.e. before the driver gene knockout, we use CID to predict the embedding *e*_1_ of masking the target gene, as well as the embedding *e*_2_ of masking both the target gene and the driver gene. We then use the gene expression classifier of scBERT to convert *e*1 and *e*2 into gene expression level *x*_1_ and *x*_2_. If *x*_1_ is less than *x*_2_, then the driver gene knockout up-regulates the target gene. Similar analogy applied to down-regulation if *x*_1_ > *x*_2_. In case *x*_1_ = *x*_2_, the driver gene has no impact on the target gene.

We obtained the up/down-regulation ground truth by comparing the scRNA-seq data of KO versus WT. DE analysis were performed for each driver gene knockout experiment, from which we classified the target genes into 3 categories: up-regulated, no-change, down-regulated. From each category, we randomly sampled 10 genes to test the performance of CID. Fig. 1D shows the predicted regulation directions versus the ground truth on the sampled target genes across all 19 experiments. It can be seen that the log2 fold change (log2fc) of target genes in the ground truth nicely agrees with CID’s predictions. This result suggests that CID can accurately predict the regulation directions without performing the actual knockout experiments.

### 3.3 CID vs CSDGI on Breast cancer data

CSDGI is a recent method that aims to find driver genes from scRNA-seq data based on ResNet. CSDGI has identified 70 driver genes in a breast cancer scRNA-seq data [13]. Here we re-analysed the scRNA-seq data using CID. First we re-trained scBERT on this data, where pre-trained weights (default setting) were used to initialize scBERT. Top 100 DE genes between caner cells and normal cells in the dataset were chosen both as the potential driver genes and target genes. Next we evaluated the marginal impact of the potential driver genes to the target genes using CID, with the impact of a gene to itself set to 0. Given the ranked driver genes by CID, we calculated the number of real driver genes by taking the top *K* ∈ {1, 2, …, 70} driver genes. If a candidate driver gene had an annotation in the NCG 7.1 database [6], we treated it as a real driver gene. Note that CSDGI only provided 70 driver genes, so we maximally take 70 potential driver genes. Fig. 1E shows the number of real driver genes identified by CID and CSDGI in the top *K* potential driver genes. It can be seen that CID identifies more annotated driver genes than CSDGI.

## 4 Conclusion

Owing to the development of LLMs, researchers could use the generated cell and gene embeddings for various causal inference based tasks. In this paper, we demonstrate CID’s superior performances in driver gene identification in three different settings. CID’s results generally offer better biological interpretations.

## Data Availability

All data produced in the present work are contained in the manuscript.

## Acknowledgments

We acknowledge the computational resources provided by the Aalto Science-IT project. This work was supported by the Research Council of Finland [grant number 335858, 358086].

## Code Availability

Code is available at https://github.com/Dionysos-o/CID/tree/master.

## References

[1] Michael S Lawrence et al. Mutational heterogeneity in cancer and the search for new cancer-associated genes. Nature, 499(7457):214–218, 2013.

[2] Abel Gonzalez-Perez and Nuria Lopez-Bigas. Functional impact bias reveals cancer drivers. Nucleic acids research, 40(21):e169–e169, 2012.

[3] David Tamborero et al. Oncodriveclust: exploiting the positional clustering of somatic mutations to identify cancer genes. Bioinformatics, 29(18):2238–2244, 2013.

[4] Uri David Akavia et al. An integrated approach to uncover drivers of cancer. Cell, 143(6):1005–1017, 2010.

[5] Evan O Paull et al. Discovering causal pathways linking genomic events to transcriptional states using tied diffusion through interacting events (tiedie). Bioinformatics, 29(21):2757–2764, 2013.

[6] Dimitra Repana et al. The network of cancer genes (ncg): a comprehensive catalogue of known and candidate cancer genes from cancer sequencing screens. Genome biology, 20:1–12, 2019.

[7] Fan Yang et al. scbert as a large-scale pretrained deep language model for cell type annotation of single-cell rna-seq data. Nature Machine Intelligence, 4(10):852–866, 2022.

[8] Haotian Cui et al. scgpt: toward building a foundation model for single-cell multi-omics using generative ai. Nature Methods, pages 1–11, 2024.

[9] F Alexander Wolf et al. Scanpy: large-scale single-cell gene expression data analysis. Genome biology, 19:1–5, 2018.

[10] Congxue Hu et al. Cellmarker 2.0: an updated database of manually curated cell markers in human/mouse and web tools based on scrna-seq data. Nucleic Acids Research, 51(D1):D870–D876, 2023.

[11] Nathan Lawlor et al. Single-cell transcriptomes identify human islet cell signatures and reveal cell-type– specific expression changes in type 2 diabetes. Genome research, 27(2):208–222, 2017.

[12] Masaya Oshima et al. Stearoyl coa desaturase is a gatekeeper that protects human beta cells against lipotoxicity and maintains their identity. Diabetologia, 63:395–409, 2020.

[13] Meng Huang et al. Unravelling cancer subtype-specific driver genes in single-cell transcriptomics data with csdgi. PLOS Computational Biology, 19(12):e1011450, 2023.

